# Projected ICU and Mortuary load due to COVID-19 in Sydney

**DOI:** 10.1101/2020.03.31.20049312

**Authors:** Andrew Francis, Yi Guo, Paul Hurley, Oliver Obst, Laurence Park, Mark Tanaka, Russell Thomson, X. Rosalind Wang

**Affiliations:** CENTRE FOR RESEARCH IN MATHEMATICS AND DATA SCIENCE, WESTERN SYDNEY UNIVERSITY; SCHOOL OF BIOTECHNOLOGY AND BIOMOLECULAR SCIENCES, UNIVERSITY OF NEW SOUTH WALES

## Abstract

The spread of COVID-19 is expected to put a large strain on many hospital resources, including ICU bed space, and mortuary capacity. In this report we study the possible demands on ICU and mortuary capacity in Sydney, Australia, using an adapted SEIR epidemiological model.

## 1. Introduction

The novel coronavirus that causes COVID-19 has been spreading rapidly throughout the world, and has emerged in several hotspots in Sydney. Because the spread in other places has been so rapid, health systems have been overwhelmed. In response, authorities have instigated a number of measures to mitigate the spread, and attempt to ease this burden.

In this study we explore the consequences of various mitigation effects on demand for critical services, in particular intensive care units (ICU), and mortuary space. The key inputs into the model are the basic reproductive number of the virus, the *R*_0_, and the effect of mitigation strategies on this reproductive number. A number of estimates of *R*_0_ for COVID-19 have now been published (e.g., [3, 5, 10]), and likewise information is being gathered about the effect of certain mitigations elsewhere on the reproductive number [5]. We use these estimates as input into a published mathematical model, and apply the model to the Sydney environment.

The model we use is an extension of the well-established SEIR family of mathematical models (Susceptible, Exposed, Infectious, Removed), that has been adapted specifically for COVID-19 by the Neher lab at the University of Basel [6]. The extension involves including various stages of illness, from infectious, to severe (in hospital), and critical (in the ICU). This level of detail is important for predicting the demands on the ICU.

Aside from running the model for a range of values of the basic reproductive number *R*_0_ (consistent with published estimated ranges) and two different mitigation effects (moderate and strong), the model takes as input a number of other key parameters. These include age-specific severity data, which we obtain from a recent Imperial College, London report [3], data on the length of stay of COVID-19 patients in each stage of hospital care, also from [3], age distribution data from Sydney [7], and a seasonal forcing effect that takes into account the likely contribution of the winter months to transmissibility.

Key findings

- For low values of the basic reproductive number, and strong effect of mitigation in Sydney, the model predicts an ICU capacity of 800 beds may be sufficient. However, for higher values of *R*_0_ or any “moderate” mitigation effect, it will not.
- The range of possible outcomes for this outbreak is vast. For example if mitigation is moderate then any of the *R*_0_ values we tested leads to ICU capacity being exceeded. At the higher end, if *R*_0_ = 2.95 then the ICU capacity is predicted to be over-run by a factor of 7.
- Demand for mortuary capacity is also very sensitive to the underlying *R*_0_ and strength of mitigation effect. For low values of *R*_0_ and strong mitigation, less than 50 spaces are predicted to be sufficient. However in any other scenario, capacity will need to exceed 150 spaces, and up to 1700 could be needed.

## 2. The extended SEIR model

We model the spread of COVID-19 in Sydney using an extended SEIR model developed by the Neher lab at the University of Basel [6]. In this model, Infectious are taken to be “in the community”, and there are several populations in the hospital system: Severe (H - in hospital), Critical (C - in ICU), and Dead (D). While normally R is “Removed”, which might include recovered and dead, we now use it for Recovered.

A schematic of the model is shown in Figure 1. The model can be expressed as a set of ordinary differential equations giving the rates of changes of the size of each sub-population. The mathematical details are given in Appendix A.

**Figure 1.**
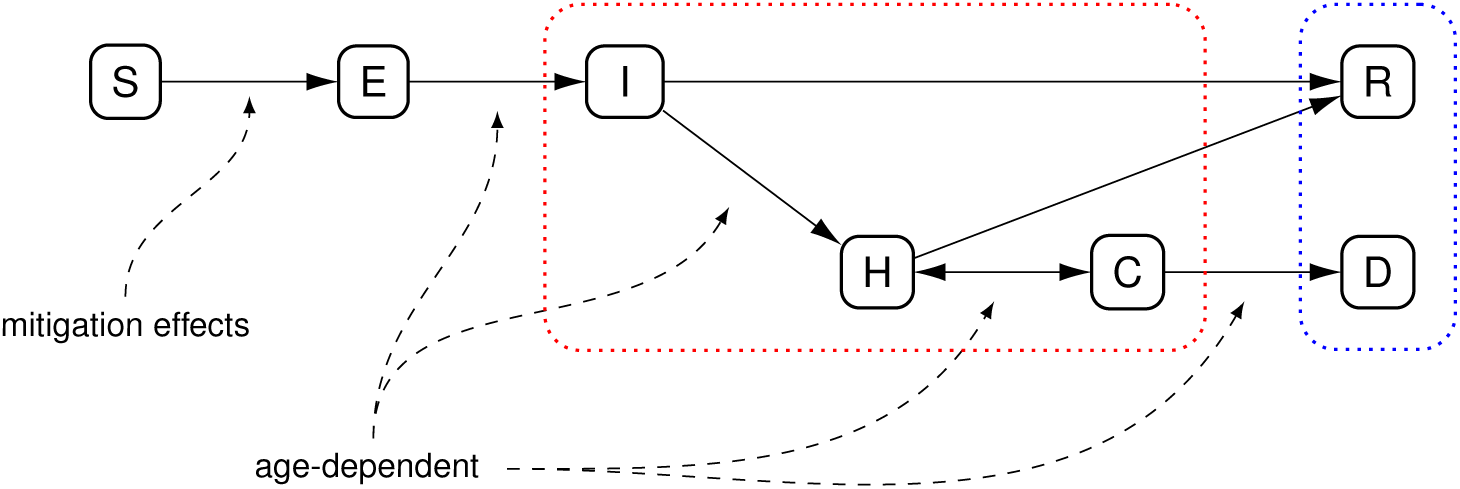
Model from the Neher lab at University of Basel [6] that we have used to generate scenarios for Sydney. The dotted red box indicates the subpopulations usually subsumed as “infectious” in traditional SEIR models, and the dotted blue box is usually treated together as “removed”.

Like other models, ours has many assumptions including the choices of parameters. We provide these details in the Appendix. The key scenarios we have modelled are:

- Basic reproductive numbers (*R*_0_) between 1.95 and 2.95.
- Strong or moderate effects of mitigation strategies (scaling the *R*_0_ by a factor of 0.45 or 0.6 respectively).

These numbers were chosen using the modeling of the team at the London School of Hygiene and Tropical Medicine, published in *The Lancet* on March 11, 2020 [5]. Their estimate of the *R*_0_ in Wuhan before any intervention is *R*_0_ = 2.35, and one week after intervention began was *R*_0_ = 1.05 (that is, 45% of 2.35). The estimate of 2.35 pre-intervention is also commensurate with the estimate of the group at Imperial College London, who arrived at a value of *R*_0_ = 2.4 [3]. Further estimates in the literature are mentioned in Appendix A. We consider the intervention in Wuhan as “strong”, and also consider a “moderate” intervention with scaling factor These factors are also close to those considered in the Neher lab modelling, which considered mitigation factors of 0.4 and 0.6.

## 3. Results

The outputs of simulations relating to hospital demand (“Severe” cases, population H) are shown in Figure 2 (a) and (b), for ICU demand (“Critical” cases, population C) are shown in Figure 2 (c) and (d), and for mortuary demand (population D) in Figure 2 (e) and (f).

**Figure 2.**
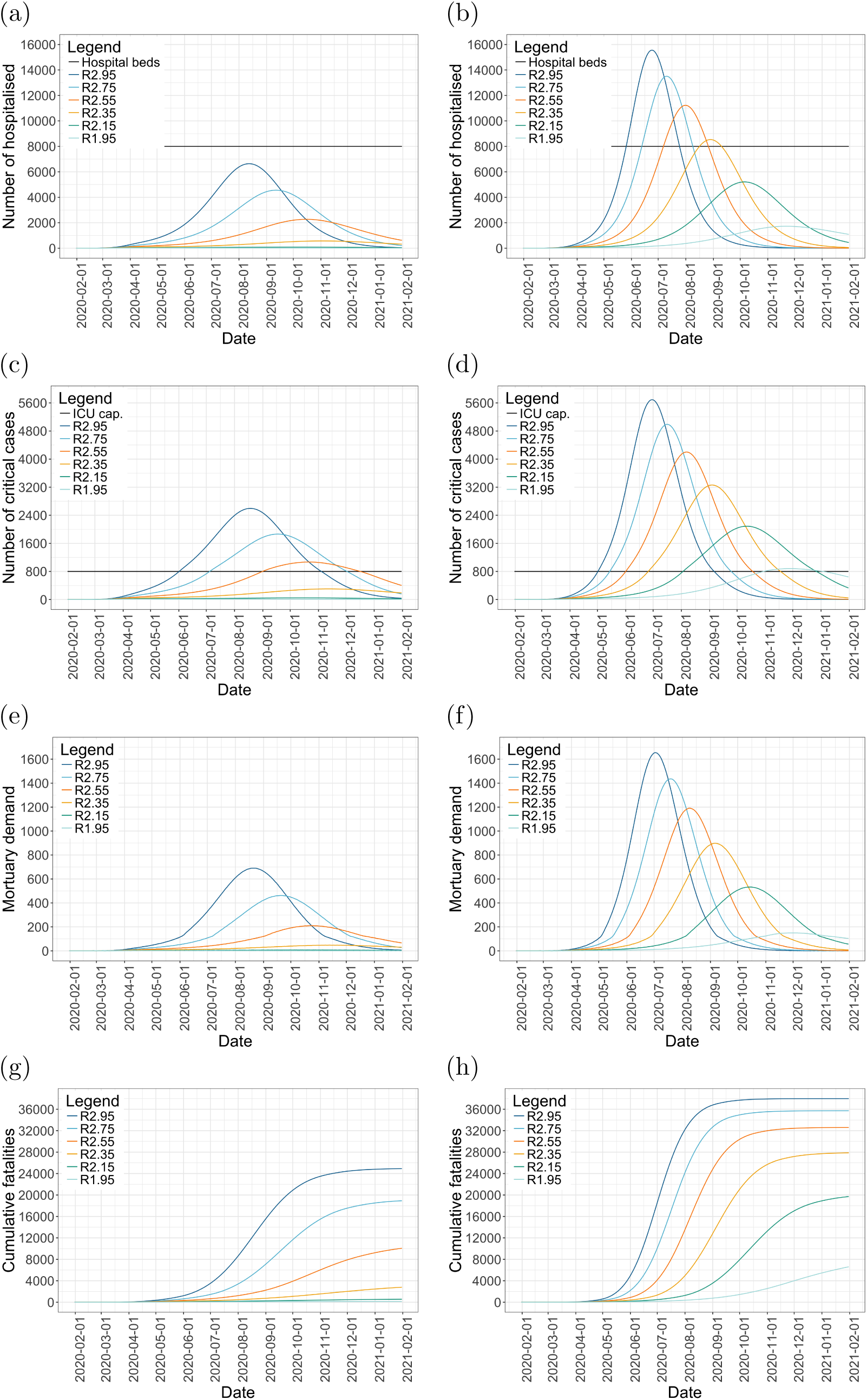
Model outputs with strong mitigation (left panels) and moderate mitigation (right panels): Hospital demand, excluding ICU, panels (a) and (b); ICU demand, panels (c) and (d); mortuary spaces required, assuming each space is occupied for 3 days, panels (e) and (f); and expected cumulative fatalities, panels (g) and (h).

**Figure 3.**
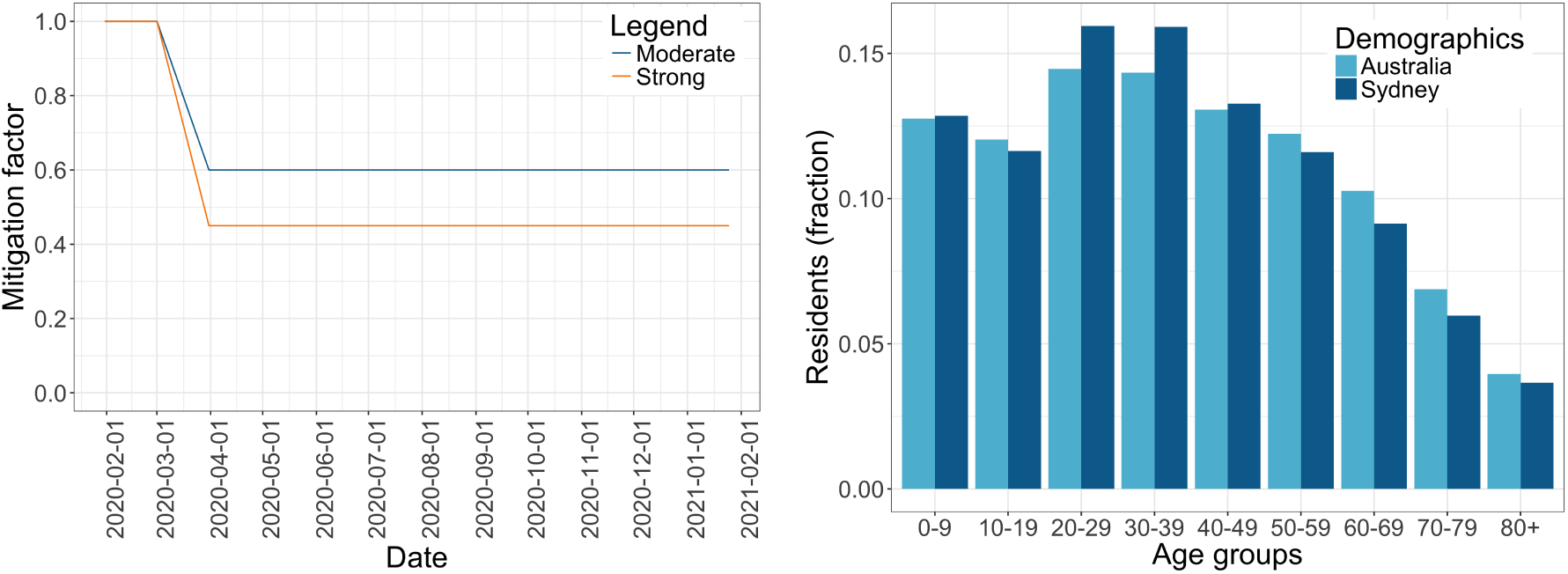
Moderate and strong mitigation strategies applied over the simulation period (left panel) are assumed to begin on March 1 2020, and be fully in effect by March 31st 2020. The demographics in the Greater Sydney region compared to the overall demographics in Australia (right panel) [7]. The younger population in Sydney compared to the rest of Australia contributes to a relatively less severe outcome of COVID-19.

Key outputs of the model are summarized in Table 1.

**Table 1.** Summary of key results, showing predicted peak values for hospital demand (excluding ICU), ICU demand, and mortuary demand, as well as the dates those peaks are expected. We also show the date the ICU capacity is predicted to be reached under each set of assumptions. The last column shows the cumulative deaths predicted under the model after one year of the simulation. A mitigation effect of *M* = 0.45 is designated “strong”, while *M* = 0.60 is “moderate”. Values of *R*_0_ = 2.35 and *M* = 0.45 (bold) are the estimated numbers for the mitigation achieved by Wuhan’s initial response. All dates in the columns are in 2020.

| Scenario |  | Peak hospital (non-ICU) demand | Date of peak hospital demand | Date ICU threshold reached | Peak ICU demand | Date of peak ICU demand | Peak mortuary demand | Date of peak mortuary demand | Cumulative deaths (Feb. 2021) |
| --- | --- | --- | --- | --- | --- | --- | --- | --- | --- |
| $R_0$ | $M$ | | | | | | | | |
| 2.95 | 0.45 | 6638 | 12 Aug | 02 Jun | 2596 | 18 Aug | 690 | 19 Aug | 24919 |
| 2.75 | 0.45 | 4559 | 11 Sep | 05 Jul | 1864 | 17 Sep | 462 | 19 Sep | 18931 |
| 2.55 | 0.45 | 2269 | 16 Oct | 01 Sep | 1068 | 20 Oct | 210 | 21 Oct | 10064 |
| **2.35** | **0.45** | **569** | **31 Oct** | — | **303** | **08 Nov** | **48** | **08 Nov** | **2798** |
| 2.15 | 0.45 | 87 | 23 Sep | — | 47 | 06 Oct | 8 | 30 Sep | 560 |
| 1.95 | 0.45 | 26 | 07 Apr | — | 13 | 12 Apr | 3 | 20 Apr | 152 |
| 2.95 | 0.6 | 15560 | 24 Jun | 02 May | 5691 | 30 Jun | 1655 | 02 Jul | 37985 |
| 2.75 | 0.6 | 13506 | 10 Jul | 15 May | 4989 | 16 Jul | 1436 | 19 Jul | 35716 |
| 2.55 | 0.6 | 11225 | 01 Aug | 01 Jun | 4201 | 07 Aug | 1191 | 09 Aug | 32596 |
| 2.35 | 0.6 | 8534 | 29 Aug | 26 Jun | 3261 | 03 Sep | 898 | 05 Sep | 27865 |
| 2.15 | 0.6 | 5207 | 06 Oct | 03 Aug | 2088 | 11 Oct | 533 | 14 Oct | 19715 |
| 1.95 | 0.6 | 1725 | 21 Nov | 30 Oct | 879 | 25 Nov | 150 | 25 Nov | 6604 |

## 4. Conclusions

The model predicts that hospital demand (excluding ICU) under the default scenario of *R*_0_ = 2.35 and strong mitigation will peak at about 570 beds by the end of October 2020. If mitigation is only of moderate effect, demand will swamp the system, with even a low *R*_0_ = 2.15 giving rise to demand for more than 5000 beds.

An ICU capacity of 800 beds is predicted to only be sufficient if there is a strong mitigation effect and for values of baseline *R*_0_ at 2.35 and below (Figure 2(c)). If the mitigation effect is only moderate (Figure 2(d)), they will not be sufficient, and there will be extreme overload beginning in May if the baseline *R*_0_ is high. Under the default *R*_0_ = 2.35 and strong mitigation, ICU demand will exceed 300 beds in November. Note: this models demand from COVID-19 alone and does not account for regular demands on the ICU capacity.

Mortuary requirements vary greatly depending on the *R*_0_ and strength of mitigation (Figure 2(e) and (f)). If *R*_0_≤2.35 and mitigation is strong, peak demand would be under 50 beds in Sydney. Any scenario with only moderate mitigation or a higher underlying *R*_0_ would result in the need for hundreds of mortuary beds.

Another output of the model is the number of fatalities over time, and this is shown in Figure 2(g) and (h). Here we see the major effect of the strength of mitigation: no scenario with only moderate mitigation predicts fewer than 6,600 fatalities. Under moderate mitigation effects, those large numbers of fatalities mostly stop increasing by late 2020, indicating the epidemic has washed through the population, albeit at massive cost to life. On the other hand, the effect of strong mitigation can be seen in the lower predicted numbers of fatalities, with some scenarios (*R*_0_ = 1.95 or 2.12) staying under 1000, but it also shows that the outbreak will persist longer, into 2021.

There are several key uncertainties in the modelling:

- What is the effect on underlying *R*_0_ of the actual mitigation strategies that have been put in place and are being considered? While it would appear that the measures introduced in Sydney — physical distancing, and the closure of non-essential businesses — should have a “strong” effect on *R*_0_, this key question is beyond the scope of this study. What we can say from the results here is that mitigation strategies need to be strong enough to produce the desired effect on *R*_0_.
- What is the baseline *R*_0_ for Sydney? The Imperial College study used 2.4 as default [3], and Kucharski *et al*. in *Lancet* estimated it at 2.35 [5]. These estimates have wide confidence intervals, and it could be lower in Sydney due to earlier awareness and also a more diffuse population. The Harvard group had a lower estimate of approx 1.9 but that assumed a quite high exposed-to-symptomatic probability of 0.5≥[10]. This assumption has the effect in their models of lowering estimates of *R*_0_.

## 5. Extensions, further questions

Some possible extensions that could be made within the same modelling framework include:

- Focusing on specific region of Sydney, which could be relevant if each hospital receives patients from local catchment.
- Including specific comorbidities may be valuable. For instance, Sydney may have higher rates of diabetes or obesity than Wuhan, which might affect death rates, while Wuhan has higher rates of smoking, especially in men. These variations could be implemented by changing the age-related parameters, assuming these conditions are aligned with different age profiles.

## Data Availability

All data in the referred to in the manuscript is publicly available; links are given in the manuscript.

https://neherlab.org/covid19/

https://itt.abs.gov.au/itt/r.jsp?databyregion#/

## Appendix A. The mathematical model and implementation

### A.1. The mathematical model

The basic model used in this study was introduced in [6], and is a generalisation of classical “SEIR” epidemiological models that have been in the literature since the SIR model was introduced in 1927 [4]. An introduction to such models can be found in the text [1].

In this implementation of the SEIR-type model shown in Figure 1, each compartment (sub-population) is further divided into age brackets, and these have different features based on what has been inferred about COVID-19 to date. Thus the model is really a set of models, one for each age bracket, denoted *a* (so that *S*_*a*_ refers to the susceptible population in age-bracket *a*). Age brackets and age-specific parameters are shown in Table 3.

**Table 2.** Parameter setting for the model.

|  |  |
| --- | --- |
| *Time parameters (taken from modelling in [3]):* |  |
| Time latent (E), before moving to I: | 6 days |
| Time infectious (I), before moving to either S or R: | 5 days |
| Time severe (S), before moving on to either C or I: | 7 days |
| Time critical (C), before moving on to either D or S: | 9 days |
| *Capacity parameters (estimates assuming doubling of ICU):* |  |
| Hospital beds Sydney: | 8000 |
| ICU beds Sydney: | 800 |
| Length of stay in mortuary (days per patient): | 3 |
| *Population parameters and initial conditions:* |  |
| Sydney population: | 5.5 mill |
| Initial infected cases*: | 50 on 1st February 2020 |
| Imports per day: | 2 |
| Seasonal forcing effect: | 0.1 |
| *Epidemiological parameters:* |  |
| $R_0$ input values (align with the range used in [3, 10]) | 1.95, 2.15, 2.35, 2.55, 2.75, 2.95 |
| Mitigation factors (reducing $R_0$): (see also Fig. 3) | 0.45, 0.6 |
| Scaling effect on rate of death given ICU needed but not available: | 2 |

\*Initial settings have been chosen because under these conditions with weak mitigation strategies, we see outbreak parameters for Sydney commensurate with those observed on 25th March.

**Table 3.**
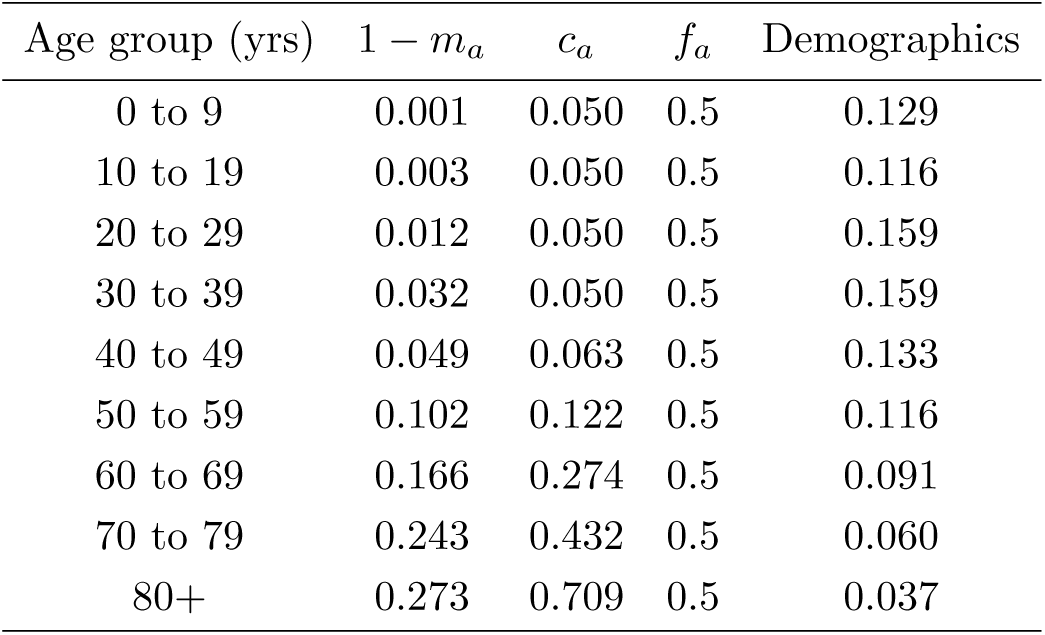
Age-specific severity parameters, from [3], and the population distribution in the Sydney region, from the Australian Bureau of Statistics [7]. Our results were computed using this age distribution specific to the Greater Sydney region (as at 30 June 2018), rather than the overall Australian distribution, with the population extrapolated to its current 5.5 million residents.

The differential equations that govern the flow of people through the compartments are as follows, as in [6]:

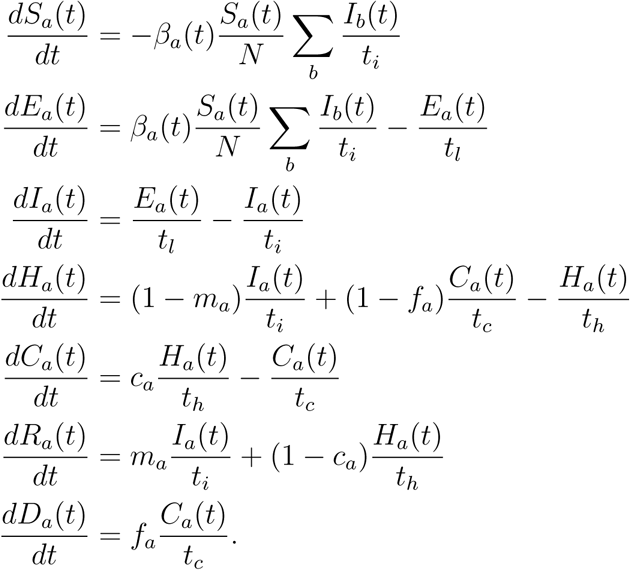

For instance, the first equation says that the rate of flow of people out from *S*_*a*_ (susceptible in age category *a*), to *E*_*a*_ (exposed) at time *t*, is the total infectious population ∑ *I*_*b*_ times the infection rate *β*_*a*_ at time *t*, times the proportion of the population still susceptible, *S*_*a*_(*t*)*/N*. Patients leave the hospital on average after *t*_*h*_ days, so each day on average *H*_*a*_(*t*)*/t*_*h*_ patients leave hospital (seen in the expression for *dH*_*a*_*/dt*). Of these, *c*_*a*_ go to the *C*_*a*_ (the ICU), and (1 −*c*_*a*_) recover (go to *R*_*a*_).

If a patient needs ICU but ICU is not available, we assume (following [6]), that leads to a fatality.

To summarize the parameters, we have:

- Transmission rate *β*_*a*_(*t*). This is essentially a function of the basic reproductive number *R*_0_, but also accounts for mitigation effects and seasonal forcing:

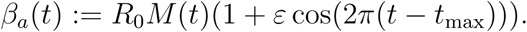 The expression for seasonal forcing is standard (see eg [1, Sect. 1.8]), but includes the mitigation effect *M* (*t*) on *R*_0_.
- Average times in each state of the model are given by
  − *t*_*l*_: latent time exposed before becoming infectious,
  − *t*_*i*_: time infectious before either recovering or becoming severely ill,
  − *t*_*h*_: time severe, in hospital, before either recovering or becoming critical,
  − *t*_*c*_: time critical, in ICU, before either dying or leaving the ICU within the hospital.
- Age-specific parameters:
  − *m*_*a*_ is the proportion of cases in *I*_*a*_ that stay mild (asymptomatic).
  − *c*_*a*_ is the proportion of cases in *H*_*a*_ that become severe.
  − *f*_*a*_ is the proportion of cases in *C*_*a*_ that are fatal.

### A.2 Parameter settings

Table 2 gives a summary of key parameter settings in our implementation of the model.

The age related parameters given in Table 3 are from [3], and originate from studies of Wuhan cases in [9].

There are now many estimates of the value of *R*_0_ in the literature, ranging from a bit below 2 to above 3. Low *R*_0_ estimates of 1.8 −2.0 were found in [10], assuming a relatively high probability that an infection becomes symptomatic of *P*_*sym*_≥0.5. Lower values of *P*_*sym*_ in that study give higher estimates of *R*_0_. An estimate of *R*_0_ = 2.2 was given in [8], using stochastic simulations and data from the early outbreak. A range of 2.0 to 2.6 was used in modelling in [3], with a default setting of 2.4. An agent-based modelling study, calibrated with similar data, estimated *R*_0_ = 2.27 [2]. Another study found pre-mitigation *R*_0_ in Wuhan to be 2.35 [5], dropping post-intervention to 1.05, which is approximately 45% of 2.35. And an estimate of *R*_0_ = 2.68 in [11], published 4th February, was based on the early spread within Wuhan and seeding of new outbreaks within China.

Taking these into account, in this study we take *R*_0_ = 2.35 as a central parameter setting (looking at several values to each side), and treat a mitigation scaling effect of 0.45 to be “strong” (since the Wuhan intervention was very strong), and a scaling of 0.6 for a moderate mitigation effect.

### A.3 Implementations of the model

The publicly available code released by the Neher lab was the starting point for our modelling, and we adapted some of the javascript to produce the outputs required for this study. In parallel, we wrote an implementation of the model in the statistical package R to give us more flexibility in extending the model and changing assumptions. The two implementations were checked for agreement on identical input parameter settings. The R package will be made available for others to check and adapt for their own use.

